# A Microbial Signature Following Bariatric Surgery is Robustly Consistent Across Multiple Cohorts

**DOI:** 10.1101/2020.11.12.20230581

**Authors:** Farnaz Fouladi, Ian M. Carroll, Thomas J. Sharpton, Emily Bulik-Sullivan, Leslie Heinberg, Kristine Steffen, Anthony A. Fodor

## Abstract

**Background:** Bariatric surgery induces significant shifts in the gut microbiota which could potentially contribute to weight loss and metabolic benefits. The aim of this study was to characterize a microbial signature following Roux-en-Y Gastric bypass (RYGB) surgery using novel and existing gut microbiota sequence data.

**Results:** We generated 16S rRNA gene and metagenomic sequences from fecal samples from patients undergoing RYGB surgery (n = 61 for 16S rRNA gene and n = 135 for metagenomics). We compared these data with three smaller publicly available 16S rRNA gene and one metagenomic datasets from patients who also underwent RYGB surgery. Mixed linear models and machine learning approaches were used to examine the presence of a common microbial signature across studies. Comparison of our new sequences with previous longitudinal studies revealed strikingly similar profiles in both fecal microbiota composition (r = 0.41 ± 0.10; p < 0.05) and metabolic pathways (r = 0.70 ± 0.05; p < 0.001) early after surgery across multiple datasets. Machine learning approaches revealed that the replicable gut microbiota signature associated with RYGB surgery could be used to discriminate pre- and post-surgical samples. Opportunistic pathogen abundance also increased post-surgery in a consistent manner across cohorts.

**Conclusion:** Our study reveals a robust microbial signature involving many commensal and pathogenic taxa and metabolic pathways early after RYGB surgery across different studies and sites. Characterization of the effects of this robust microbial signature on outcomes of bariatric surgery could provide insights into the development of microbiome-based interventions for predicting or improving outcomes following surgery.

**Trial Registration:** clinicaltrials.gov, number NCT03065426. Registered 27 February 2017, https://www.clinicaltrials.gov/ct2/show/NCT03065426

## Introduction

Bariatric surgery, the most effective treatment for severe obesity, induces rapid and durable weight loss and improves or resolves many obesity-associated comorbidities including Type II diabetes and cardiovascular diseases [1]. Mechanisms that have been proposed to contribute to weight loss and metabolic improvements following bariatric surgery include changes in diet, hormones, bile acids, energy metabolism, and the gut microbiome [2]. Recent literature has shown that the composition and function of the gut microbiome undergo significant changes following bariatric surgery and this may partly contribute to its beneficial effects; however, the underlying mechanisms for such effects remain unknown [3-5].

To clarify potential mechanisms by which the gut microbiome exerts its beneficial effects, several studies have evaluated how bariatric surgery alters the gut microbiome. While these studies identify taxa that change in response to surgery, the specific microbiome alterations reported across studies have been inconsistent. For example, both increases and decreases in the relative abundance of the phylum *Bacteroidetes* as well as *Faecalibacterium* and *Bifidobacterium* species following bariatric surgery have been reported [3, 6-13]. These differences can possibly be explained by variation in study design between research groups (e.g. demographics, geography, exclusion and inclusion criteria, sample size, and dietary intake) or differences in laboratory techniques (e.g. DNA extraction, and sequencing platform) [14]. In areas such as obesity and colorectal cancer, prior meta-analyses have integrated microbiome data across multiple studies to resolve robust associations between the gut microbiome and health covariates [15-19]. Such studies apply consistent bioinformatic tools and statistical models to all datasets to eliminate analytical variability between investigations and, where possible, model the impact of study design and methodological factors on the results. However, no such meta-analysis has yet clarified whether consistent associations exist between changes in the gut microbiome and RYGB surgery outcomes across studies.

In this study, we generated 16S rRNA gene and metagenomics data from fecal samples obtained from patients who underwent Roux-en-Y Gastric Bypass (RYGB) surgery and compared the microbial signature observed from our study with three publicly available 16S rRNA gene datasets and one metagenomic dataset obtained from similar RYGB-microbiota studies while controlling for sequencing and bioinformatic tools. Our study reveals that much of the microbial community of patients in different cohorts responds to surgery in very similar ways. Establishment of this consistent microbial signature is a necessary pre-requisite for development of robust clinical tools that could use microbial changes to predict personalized outcomes to bariatric surgery.

## Methods

### Publicly available 16S rRNA gene and metagenomic datasets

In this study, we generated novel 16S rRNA gene and metagenomic datasets from fecal samples from patients undergoing RYGB and performed a comparative analysis of changes in the fecal microbiota from pre-to-post RYGB using our novel dataset and previously published datasets. For this, we searched NCBI for Bioprojects that included clinical studies involving patients undergoing RYGB surgery as well as their associated 16S rRNA gene sequences, metagenomics and metadata (**Supplementary Table 1**). Studies that included longitudinal samples and were sequenced on the Illumina platform were included in our comparative analysis, while cross-sectional studies were excluded. Three 16S rRNA gene datasets and one metagenomic dataset met these requirements [12, 20-22]. The raw sequences for these studies were obtained from the Sequence Read Archive (SRA) with project numbers specified in **Table 1**. The dataset from the Afshar et al. study was part of Biomarkers Of Colorectal cancer After Bariatric Surgery study (www.isrctn.com/ISRCTN95459522) and no publication for the microbiome dataset is available. Also, for this dataset, the time for collection of post-RYGB samples is not determined in the metadata, however, based on Afshar et al 2018 [22], patients were followed up at 6 months after surgery. The Ilhan et al 2020 study included mucosal samples from post-RYGB patients and fecal samples from a cross sectional cohort; however, these samples were removed from our analysis. Sample collection, DNA extraction, and library preparation can be found in original publications and have been summarized in Supplementary Table 1.

**Table 1.** Patient Characteristics and number of samples before and after Roux-en-Y Gastric Bypass Surgery. N indicates the total number of samples for each dataset. ^a^ Four samples at three years after RYGB from the Assal study are not included in the downstream analysis. ^b^ Patient characteristics were not available in the associated metadata. ^c^ The Afshar study was part of Biomarkers of Colorectal cancer After Bariatric Surgery study (http://www.isrctn.com/ISRCTN95459522). No publication for this microbiome dataset is available. The time for the collection of post-RYGB samples was not specified in the metadata, however, based on Afshar et al 2018 patients were followed up at six months after surgery. Age and BMI (kg/m^2^) are presented as mean ± standard deviation.

| Project # | Sequence type | Total # of samples | Pre-surgery | Post-surgery 1 month | Post-surgery 3 months | Post-surgery 6 months | Post-surgery 1 year | Post-surgery 2 years | Post-surgery 3 years | Publication | Dataset names | Age | BMI | Sex (F/M) | Site |
| --- | --- | --- | --- | --- | --- | --- | --- | --- | --- | --- | --- | --- | --- | --- | --- |
| PRJNA612042 | 16S rRNA gene | 22 | 9 | -- | -- | 7 | 6 | -- | -- | Ilhan et al 2020 | Ilhan | --<sup>b</sup> | --<sup>b</sup> | --<sup>b</sup> | Arizona, USA |
| PRJNA395660 | 16S rRNA gene | 71 | 25 | -- | 20 | -- | 14 | 8 | 4<sup>a</sup> | Assal et al 2019 | Assal | 45.8 ± 7.1 | 46.4 ± 5.5 | 25/25 | Sao Paulo, Brazil |
| PRJEB14082 | 16S rRNA gene | 38 | 19 | -- | -- | 19<sup>c</sup> | -- | -- | -- | BOCABS Afshar et al 2018 | Afshar<sup>c</sup> | --<sup>b</sup> | --<sup>b</sup> | --<sup>b</sup> | UK |
| PRJEB12947 | WGS | 33 | 13 | -- | 12 | -- | 33 | -- | -- | Palleja et al 2016 | Palleja | --<sup>b</sup> | --<sup>b</sup> | 8/5 | Denmark |
| This study | 16S rRNA gene | 61 | 30 | 21 | -- | 10 | -- | -- | -- | --- | BS | 43.5 ± 10.0 | 44.7 ± 7.1 | 26/4 | Fargo and Ohio, USA |
| This study | WGS | 135 | 52 | 38 | -- | 27 | 18 | -- | -- | --- | BS | 42.8 ± 9.8 | 45.1 ± 6.1 | 46/6 | Fargo and Ohio, USA |

### Clinical study (BS dataset)

We used data (BS dataset, **Table 1**) from our ongoing prospective study, which aims to examine the impact of biological and behavioral variables on weight outcomes following bariatric surgery [23]. This study is registered at www.clinicaltrials.gov with trial ID NCT03065426. Details regarding specific aims and study design for this study have been previously published [23]. Briefly, patients are recruited at two sites: Sanford Center for Biobehavioral Research (CBR), in collaboration with North Dakota State University, in Fargo, ND, and Cleveland Clinic Bariatric and Metabolic Institute, Cleveland, OH. This study was approved by the Institutional Review Boards (Cleveland Clinic: #16-1460, Sanford Health: #00001409 and North Dakota State University #PH17112) at both data collection sites and all patients provided written informed consent prior to enrollment. Female or male patients, aged 18-65 years undergoing a first bariatric surgery procedure, either a RYGB or Sleeve Gastrectomy (SG), were recruited and evaluated at baseline (pre-surgery) and multiple timepoints (one, six, 12, 18, and 24 months) following surgery. Exclusion criteria can be found in Heinberg et al 2020 [23]. For this study, samples at baseline, one, six, and 12 months post-RYGB were included as samples at 18 and 24 months are not yet available at the time of preparing this manuscript.

### Microbiome analysis (BS dataset): 16S rRNA gene and metagenomics

Fecal samples were collected at baseline and each timepoint after surgery and stored at −80 °C until analysis. DNA extraction was performed as previously described [24]. Briefly, a phenol/choloroform extraction method combined with physical disruption of bacterial cells and a DNA clean-up kit (Qiagen DNeasy Blood and Tissue extraction kit, Valencia, CA) was used to extract DNA from fecal samples. For 16S rRNA gene sequencing, the V4 variable region was amplified by polymerase chain reaction (PCR) using 16S rRNA gene primers (forward: 5’-CAACGCGARGAACCTTACC-3’; reverse: 5’-CAACACGAGCTGACGAC-3’) and sequenced on the Illumina MiSeq platform (Illumina, San Diego, CA) at the High-Throughput Sequencing Facility in the Carolina Center for Genome Sciences at the UNC School of Medicine as previously described [25]. For metagenomics, extracted DNA was subjected to 2×150 bp paired-end sequencing on the Illumina HiSeq 4000 platform at the UNC-Chapel Hill high throughput sequencing facility.

### Comparative, integrated analysis across multiple datasets

Forward reads from the four 16S rRNA gene datasets were individually run through the DADA2 pipeline [26] to generate amplicon sequence variants (ASVs). Sequences were filtered using the “filterAndTrim” function in DADA2 with default parameters and maxEE = 2 (reads with expected errors more than 2 were discarded). For the BS, Assal, and Afshar datasets forward reads were truncated at 200 base pair and for Ilhan study reads were truncated at 150 base pairs. The SILVA132 database was used for taxonomic classification of ASVs using the DADA2 “assignTaxonomy” function. The metagenomic datasets (Palleja and BS) sequences were analyzed with Kraken2 [27] using the BioLockJ automated pipeline (https://github.com/BioLockJ-Dev-Team/BioLockJ). In order to compare changes in the abundance of opportunistic pathogens at the species level after surgery across cohorts, 16S rRNA gene datasets were taxonomically classified using the Kraken2 pipeline in addition to the metagenomic datasets. Recent work by Lu et al 2020 [28] showed that classification of 16S rRNA gene microbial communities using Kraken2 is fast and accurate.

Each 16S rRNA gene and metagenomic dataset (from DADA2 or Kraken2) was normalized using the following formula to correct for different sequence depth across samples [29]:

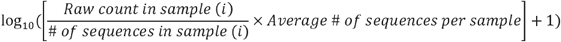

Metabolic pathways for the metagenomic datasets were profiled using the HUMAnN2 pipeline [30] and the MetaCyc database [31]. Pathway abundances (reads per kilobase) were normalized to copies per million using the “humann2_renorm_table” function from HUMAnN2.

For each dataset, longitudinal fecal samples from patients who underwent RYGB were selected. Samples from other procedures such as SG or cross-sectional samples were removed from the downstream analysis. Taxa that were present in less than 10% of samples were removed. Mixed linear models with patients as a random effect and timepoint as a fixed effect (taxa ∼ timepoint, random = ∼1 | patient ID) were constructed to compare the log_10_ normalized count of taxa between pre-surgery and different timepoints post-surgery using the “lme” function from the nlme package in R. P-values were generated from the “summary” function in R for each post-surgery timepoint with a baseline reference. Mixed linear models were used rather than paired t-tests because not every patient had samples for all timepoints (**Table 1**). To compare results across studies, -log_10_ unadjusted p-values from mixed linear models were multiplied by the sign of the regression slope and “log_10_ p-values vs. log_10_ p-values” plots were generated. Therefore, positive and negative log_10_ p-values indicate an increase and decrease, respectively, in log_10_ normalized count of taxa after surgery compared to baseline. Spearman’s rank-order correlation was used to examine the correlations of the log_10_ p-values between studies or between different timepoints within a study. P-values from the Spearman’s rank-order correlations were corrected for multiple hypothesis testing using the Benjamini–Hochberg procedure and were considered to be significant when the False Discovery Rate (FDR) was <5%. In addition, a hierarchical clustering heatmap based on the Euclidian distances of the log_10_ p-values from mixed linear models were generated using the function “Heatmap” from the package ComplexHeatmap in R. Similar statistical analyses were performed to compare changes in metabolic pathways post-surgery between the BS and Palleja cohorts.

Principal coordinate analysis with Bray-Curtis distance was performed on log_10_ normalized count of taxa from the joined four 16S rRNA gene sequencing datasets using the “capscale” function from the Vegan package in R. The PERMANOVA test on Bray-Curtis distances was further used to compare the microbiota composition between different datasets and timepoints (pre-versus post-surgery) after surgery (Microbiota ∼ study * timepoint). Shannon diversity index, a measurement of richness and evenness, was performed using the “diversity” function from the Vegan package in R. Finally, supervised classification models, including Random Forest and Least Absolute Shrinkage and Selection Operator (LASSO) were used to examine the microbial signature following RYGB surgery. For this purpose, we used a statistical workflow developed by Wirbel et al 2019 [15]. In this workflow, taxa at the genus level that have no variance across samples are removed and then are log-transformed after adding a pseudocount of 1 × 10−5 and standardized as z-scores. Each dataset was split into a train set and a test set with 8-fold cross validation and 10 repeats. Within study prediction of surgical status (pre-surgery versus post-surgery) in a test set was calculated as average area under the receiver operating characteristic curve (AUROC) from 10 trained models (one model from the 8-fold cross-validation * 10 repeats). Additionally, the trained models from all cross validations and repeats (180 models: 8 models from cross validation * 10 repeats) were then used to predict the surgical status in another dataset. Also, all joined datasets except for one dataset were trained to predict the hold-out dataset (leave-one-study-out (LOSO) validation). Predictions were averaged across all models. All analyses and visualizations were performed using R studio (Version 1.3.1056).

## Results

### Inference using mixed linear models reveals a consistent fecal microbiota signature associated with RYGB surgery across multiple 16S rRNA gene datasets

We generated 16S rRNA gene sequence data from fecal samples obtained from patients before (n=30) and one month (n=21) and six months (n=10) after RYGB surgery. We combined these data with three smaller publicly available 16S rRNA gene sequence datasets from patients who also underwent RYGB surgery. Demographics and number of samples at each timepoint are presented in Table 1. Consistent with many previous studies showing large differences between cohorts analyzed by different groups [32], PCoA ordination using Bray-Curtis distance showed pronounced clustering by study (**Figure 1A**). Using the PERMANOVA test with independent variables of study, time (pre-versus post-surgery) and the interaction between time and study showed highly significant differences between the gut microbiota compositions across studies (R^2^ = 0.33, p = 0.001, **Figure 1A**). Although the effect size was much smaller, we also observed a significant difference between pre- and post-surgical samples (R^2^ = 0.03, p = 0.001, **Figure 1B**), while the interaction between study and time was not significant (R^2^ = 0.01, p = 0.22). No significant differences in Shannon Diversity Index was observed between different timepoints (**Supplementary Figure 1**).

**Figure 1.**
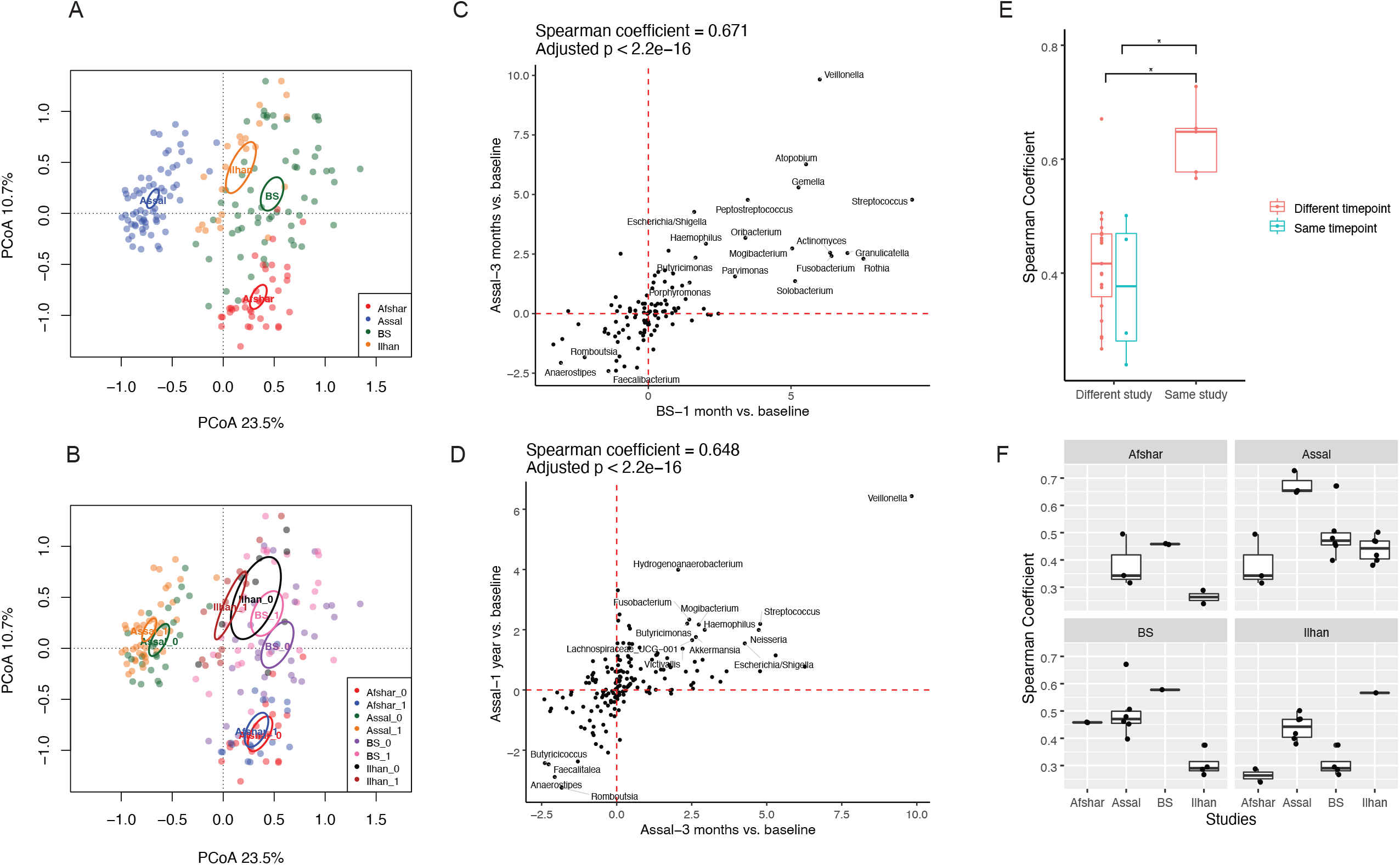
A Consistent Signature of Fecal Microbiota at the Genus Level is Associated with Roux-en-Y Gastric Bypass Surgery Across 16S rRNA gene Studies. (A) Principal coordinate analysis using Bray-Curtis distance was performed on the fecal microbiota composition at the genus level. Samples are clustered by study, suggesting significant differences present across studies. (B) A similar ordination plot to A is shown with pre-surgical samples (0) and post-surgical samples (1) specified for each dataset. (C & D) log_10_ p-value versus log_10_ p-value plots are generated using the unadjusted p-values from mixed linear models comparing log_10_ normalized count of taxa at each timepoint compared to baseline with patient ID as random effects. Upper right-quadrant and lower left-quadrant show taxa that were increased and decreased, respectively, in two different studies (C) or at two different timepoints within a study (D). Spearman rank-order correlation was used to test the consistency of changes in taxa after RYGB between studies or within studies. Taxa with unadjusted p < 0.05 in both studies or at both timepoints within a study are annotated. All pairwise comparisons between and within studies are shown in Supplementary Figure 2. (E) Boxplots show the correlation coefficients from Spearman rank-order correlations between and within studies. Wilcoxon signed-rank test was used to compare the correlation coefficients (* adjusted p < 0.05). Post-surgery time for Afshar study was assumed to be 6 months based on Afshar et al 2018. (F) Boxplots show correlation coefficients between different studies or between different timepoints in a study.

In order to provide a more detailed view of taxa that were changing with surgery, for each study we compared the log_10_-normalized count of each genus at each timepoint after surgery to the same genus’s normalized baseline abundance using mixed linear models. To enable comparison of results across studies, we created “log_10_ p-value vs. log_10_ p-value” plots in which the results from the “pre-vs-post” coefficients of our models at each timepoint were compared across pairs of studies (**supplementary Figure 2**). For example, **Figure 1C** compares the changes in genera from baseline to post-surgery between our BS study dataset at one month and the Assal study at three months post-surgery, which had the greatest similarity across all pairs of studies. Genera in the upper right quadrant are enhanced post-surgery in both studies while genera in the lower left-quadrant are reduced post-surgery in both studies and genera with unadjusted p < 0.05 in both studies are annotated. Using this comparison, there is a remarkable degree of similarity (adjusted p < 0.001 spearman r = 0.671) across the two studies. We made similar plots comparing different timepoints for the same studies (**Figure 1D** and **Supplementary Figure 2**) in which we compared changes in the log_10_-normalized count of genera at each timepoint compared to baseline and then showed the consistency between those comparisons using “log_10_ p-value vs. log_10_ p-value” plots. All five pairwise comparisons within studies and the 23 pairwise comparisons across studies were significantly associated at a 5% FDR, although as we would expect from our PCoA analysis, there was greater consistency comparing timepoints within studies than across studies (r = 0.63 ± 0.07 versus r = 0.41 ± 0.10) (**Figure 1E**). Among the four 16S rRNA gene datasets, the Ilhan study was somewhat of an outlier exhibiting weaker associations with the Afshar and our novel BS study **(Figure 1F)**. The Ilhan study had the smallest sample size among the datasets (**Table 1**) and this may explain why its associations with other studies were somewhat less robust.

Since the DADA2 pipeline allowed us to increase the resolution of classification of 16S rRNA gene sequences to sequence variants, we explored whether the high degree of similarity between studies is present at the high-resolution sequence variant level. For this, we compared changes in the log_10_ relative abundance of sequence variants between the BS and Assal studies as these studies included amplicons from the same region of the 16S rRNA gene with the same length and primers. We observed that changes in sequence variants post-surgery followed a similar pattern in both studies (**Supplementary Figure 3**) and that in some cases sequence variants that were increased or decreased in both studies belonged to different genera, such *as Veillonella, Atopobium, Fusobacterium, Lachnoclostridium*, and *Streptococcus* (**Supplementary Figure 4**). Overall, we conclude that the genus and sequence variant view give highly concordant results in the comparison of these two datasets.

Following our observation of significant correlations between studies, we next determined which genera were consistently increased or decreased across studies compared to baseline. We observed that changes in the relative abundance of *Veillonella, Streptococcus, Gemella, Fusobacterium, Escherichia/Shigella*, and *Akkermansia* showed a similar trend and increased at least at one timepoint after surgery across the four 16S rRNA gene datasets. *Rothia, Actinomyces, Atopobium*, and *Granulicatella* showed a similar trend in changing after surgery across three datasets, but not in the Ilhan study. On the other hand, *Blautia* decreased at all the timepoints after surgery in all four 16S rRNA gene datasets (**Figure 2**). As most significant changes relative to baseline are observed at one month post-surgery in the BS study and three months post-surgery in the Assal study, these timepoints from these two studies clustered together using hierarchal clustering based on Euclidian distances (**Figure 2)**. These results may reflect greater shifts in the microbiota during the first few months after surgery or may be simply explained by the larger sample size at these timepoints for both the BS and Assal studies.

**Figure 2.**
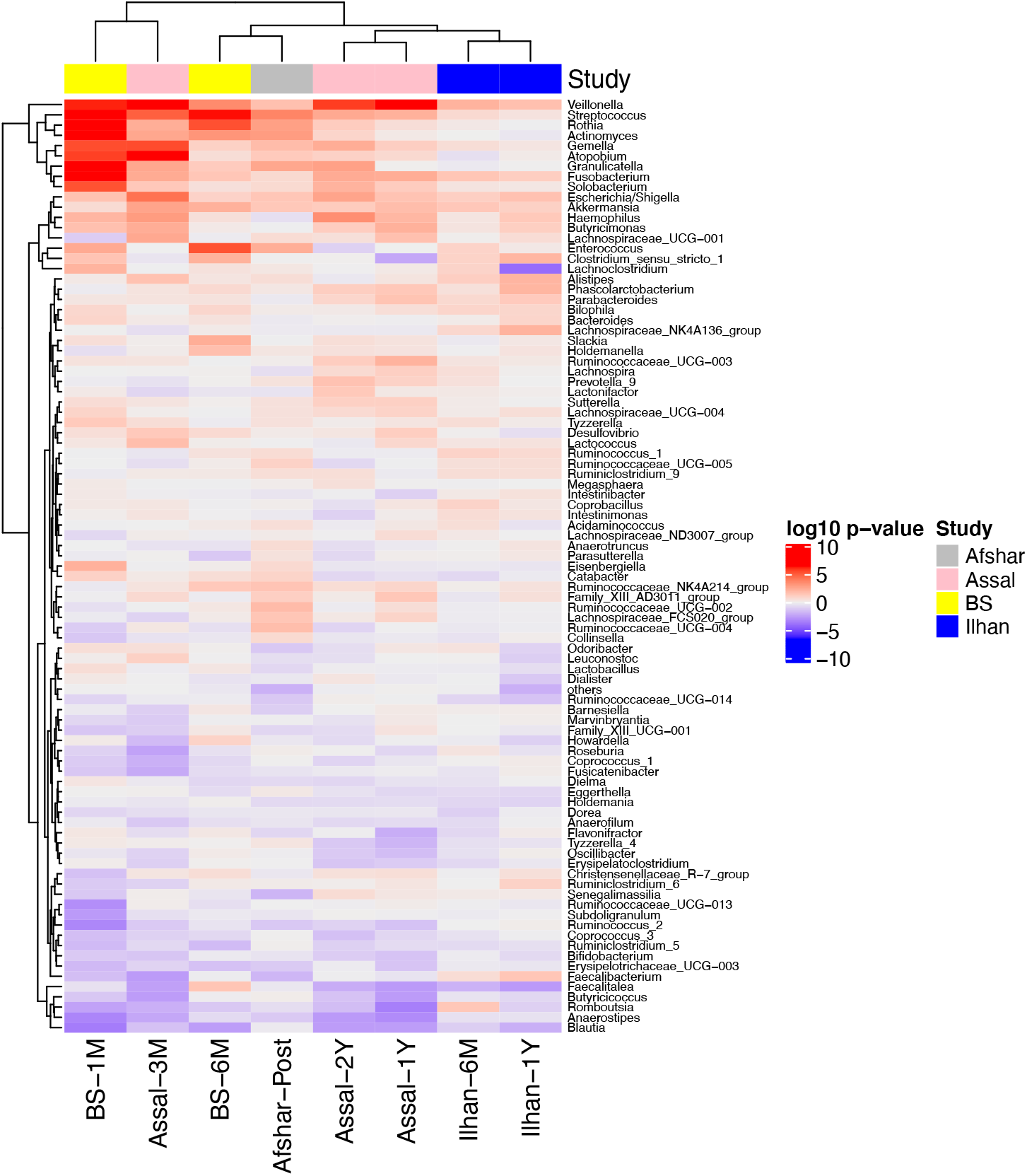
Several Taxa at the Genus Level were Increased or Decreased Consistently Across Datasets. A hierarchical clustering heatmap was performed on the Euclidian distances of log_10_ p-values from the mixed linear models with timepoint as a fixed effect and patient ID as a random effect. Red indicates taxa were increased after surgery, while blue indicates taxa were decreased after surgery compared to baseline.

### Metagenomic studies reveal a consistent and robust signature in microbial pathways associated with RYGB surgery across studies

The above results relied on the SILVA database contextualized taxonomic annotations that stemmed from analysis of DADA2 clustered 16S rRNA gene sequences. In order to show that these results are independent of the database used for taxonomic classification as well as the 16S rRNA gene sequencing platform, we performed metagenomics on 135 samples from the BS study, taxonomically classified sequences using the Kraken2 classifier, and compared changes in the log_10_ relative abundance of species at each timepoint relative to baseline with a smaller publicly available metagenomic dataset [12] (Palleja, Table1). Similar to the results from pairwise comparison of 16S rRNA gene datasets, a significant correlation was observed when we compared the changes in species from baseline to post-surgery between our BS study and the Palleja study (r = 0.31 ± 0.04, p < 0.001; **Figure 3A and Supplementary Figure 5**), suggesting some taxa have similar patterns of changing post-surgery in both studies. For example, *Streptococcus spp*., *Klebsiella spp*., *Veillonella parvula, Citrobacter freundii*, and *Enterobacter hormaechei* were increased in both studies. As expected, a greater consistency in changes in species was observed when we compared different timepoints within the Palleja or BS studies (r = 0.76 ± 0.06 versus r = 0.31 ± 0.04; **Figure 3B and 3C and Supplementary Figure 5)**. While significant, the correlation based on taxa from these metagenomic studies was smaller than we observed for 16S rRNA gene studies.

**Figure 3.**
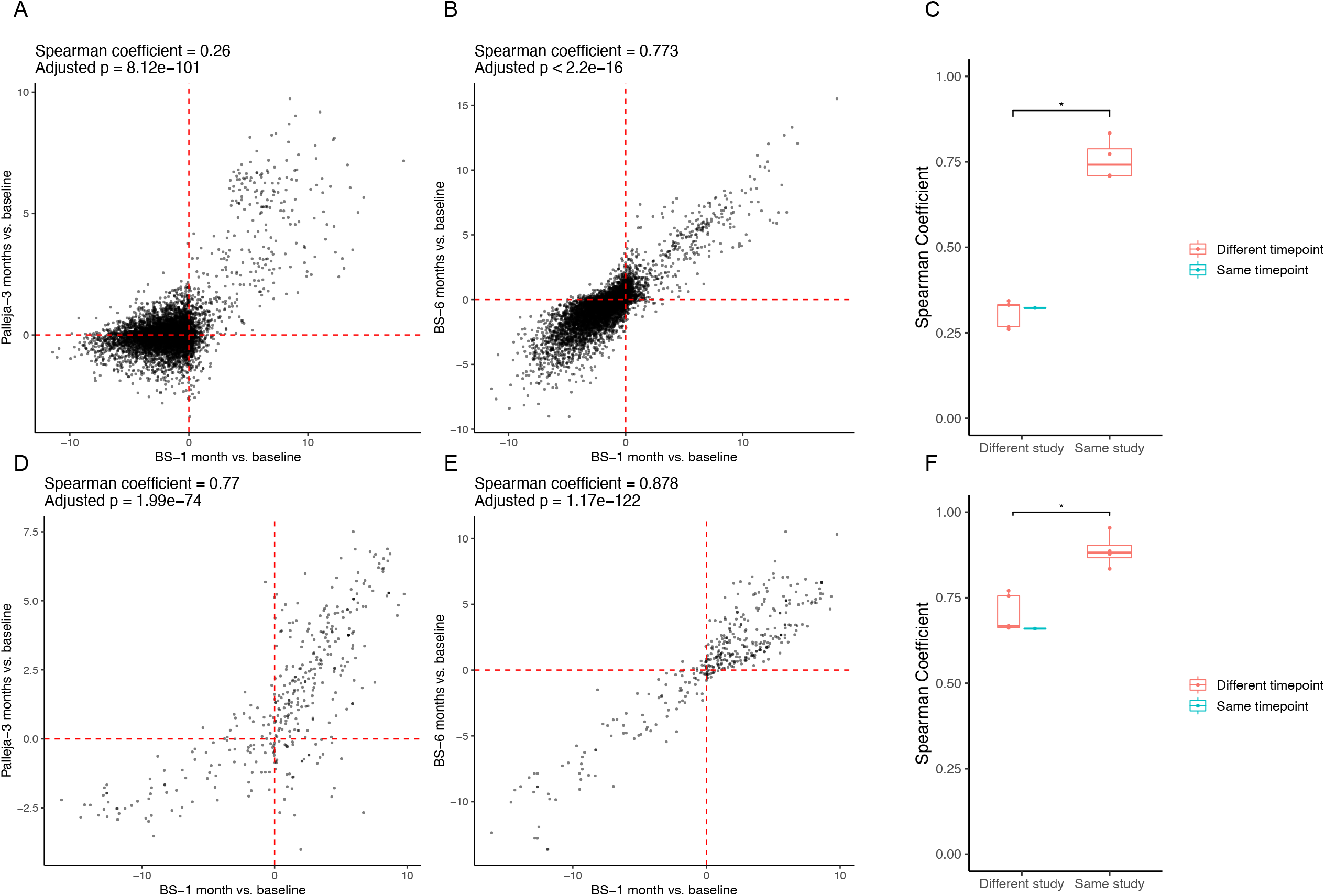
A Consistent Signature of Metabolic Pathways in Addition to Microbiota Composition is Associated with Roux-en-Y Gastric Bypass Surgery Across Metagenomic Studies. (A-B) log_10_ p-value versus log_10_ p-value plots are generated using the unadjusted p- values from mixed linear models comparing log_10_ normalized count of species at each timepoint post-surgery to baseline. The upper right-quadrant and lower left-quadrant show species that were increased and decreased, respectively, in two different studies (A) or at two different timepoints within a study (B). Spearman rank-order correlation was used to test the consistency of changes in species after RYGB between studies or within studies. All pairwise comparisons between and within studies are shown in Supplementary Figure 5. (C) Boxplots show the correlation coefficients from Spearman rank-order correlations between and within studies. Wilcoxon signed-rank test was used to compare the correlation coefficients (* adjusted p < 0.05). (D-F) Similar plots to (A-C) are generated but for metabolic pathways profiled by the HUMAnN2 pipeline.

In addition, changes in abundances of metabolic pathways profiled by the HUMAnN2 pipeline were compared between these two metagenomic datasets. Interestingly, compared to the taxonomic composition, changes in abundances of metabolic pathways post-surgery relative to baseline showed a greater degree of similarity between the Palleja and BS datasets (r = 0.70 ± 0.05; p < 0.001 across different timepoints between studies; r = 0.89 ± 0.05; p < 0.001 across different timepoints within studies; **Figure 3D-F and Supplementary Figure 6**). Notably, pathways involving gluconeogenesis, L-alanine biosynthesis, biotin biosynthesis, hexitol degradation, mycolate biosynthesis, N-acetylneuraminate degradation, and fatty acid elongation were enriched post-surgery in both studies, while dTDP-L-rhamnose biosynthesis I and starch degradation were decreased pos-surgery in both studies (**Supplementary Table 2)**. These results suggest that functional microbial genes respond similarly to RYGB surgery across cohorts.

### Machine learning approaches confirm that the fecal microbiota can be used to discriminate pre- and post-surgery samples

Machine learning approaches have often been used in metagenomic analyses to determine whether a microbial signature in one study can be used to predict the phenotype of samples from another study [33]. In order to determine if such an approach could be extended to our analyses, we next used supervised classifiers to assess whether the signature of the gut microbiota after surgery generalizes across 16S rRNA gene studies. For this purpose, we used the Least Absolute Shrinkage and Selection Operator (LASSO) logistic regression (**Figure 4**) and Random Forest (**Supplementary Figure 7**) classifiers implemented in a statistical workflow developed by Wirbel et al 2019 [15]. Within study cross validation performance, quantified by the area under the receiver operating characteristic curve (AUROC) was highest for the BS and Assal studies (above 0.90 for both LASSO and Random Forest classifiers) and was the lowest for the Ilhan study (Lasso: 0.67, Random Forest: 0.59) and Afshar study (LASSO: 0.67, Random Forest: 0.74) (**Figure 4A and Supplementary Figure 7A**), which likely reflect the smaller sample size of these latter studies. The average study-to-study accuracy predictions ranged between 0.66 and 0.84 for the LASSO classifier with the highest performance being associated with using Afshar or BS studies as training sets (0.84 and 0.83, respectively) and the lowest performance associated with training on the Ilhan study (0.66) (**Figure 4A)**. Finally, we examined when a classifier is trained using all datasets but one, how well it generalizes in evaluation on the remaining hold-out study (leave-one-study-out: LOSO validation). The LOSO performance for all datasets was between 0.77-0.95 using the LASSO classifier (**Figure 4B)** and was above 0.85 using the Random Forest classifier (**Supplementary Figure 7B**). All these results are consistent with the RYGB procedure having a replicable microbiota signature across studies.

**Figure 4.**
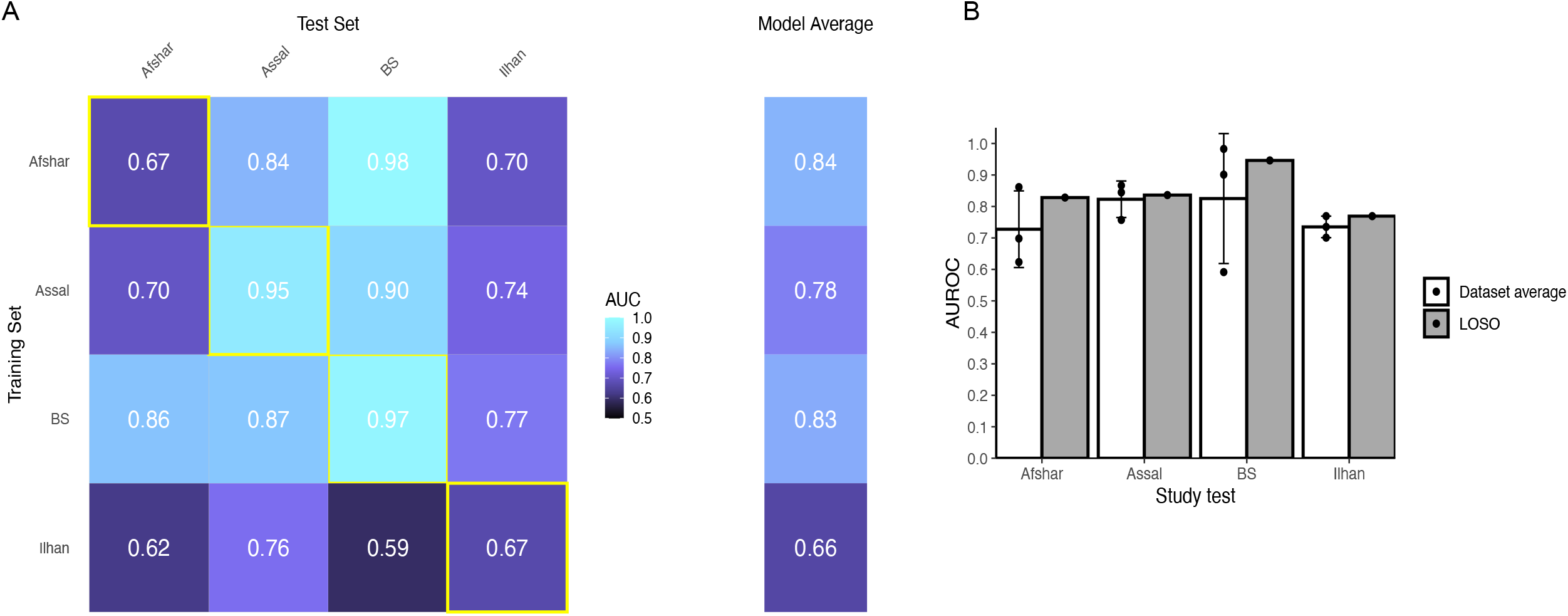
Supervised Classification Using the Least Absolute Shrinkage and Selection Operator Logistic Regression Reveals that the Fecal Microbiota can be Used to Discriminate Pre- and Post-Samples Across 16S rRNA gene Datasets. (A) The heatmap shows the area under the receiver operating characteristic curve (AUROC) from cross validations within each study (yellow boxes along diagonal) and study-to-study model transfer (external-validations off-diagonal). The last column shows the average AUROC for study-to-study predictions. (B) White bar plots show the AURC for a model trained on data from a single study predicting the test study on the X-axis. The bar height represents the average AURC for the 3 classifiers and the error bar represents the standard deviation. The grey bar plots show the AURC for a model trained on all studies but one (LOSO validation).

### Abundances of opportunistic pathogen increase post-surgery in a consistent manner across cohorts

Since we observed similar changes in the microbial community following surgery across studies, we next asked whether changes in the gastrointestinal environment post-surgery provide an opportunity for opportunistic pathogens that could be acquired from the hospital or could be associated with antibiotic treatment to flourish in the gastrointestinal tract. For this purpose, from our analysis from the Kraken2 pipeline, for both 16S rRNA gene and metagenomic sequences, we compared the log_10_ normalized count of typical opportunistic pathogens *Klebsiella pneumoniae, Klebsiella oxytoca, Enterococcus faecalis, Enterobacter cloacae complex sp, Enterobacter cloacae, Enterococcus faecium*, and *Clostridium perfringens* between different timepoints using a mixed linear model for each study. Interestingly, our BS metagenomic and Palleja metagenomic datasets showed a largely consistent set of opportunistic pathogens that increased at an adjusted p < 0.05 one month or three months post-surgery and remained high in abundance at later timepoints following surgery (**Figures 5 A and B**). One exception to this agreement between studies was *Enterococcus faecium* which decreased in the BS study following surgery but was unchanged in the Palleja study. In addition, an increase in *Clostridium perfringens* reached statistical significance (adjusted p = 0.001) at six months post-surgery in our BS metagenomic study but not in the Palleja metagenomic study.

**Figure 5.**
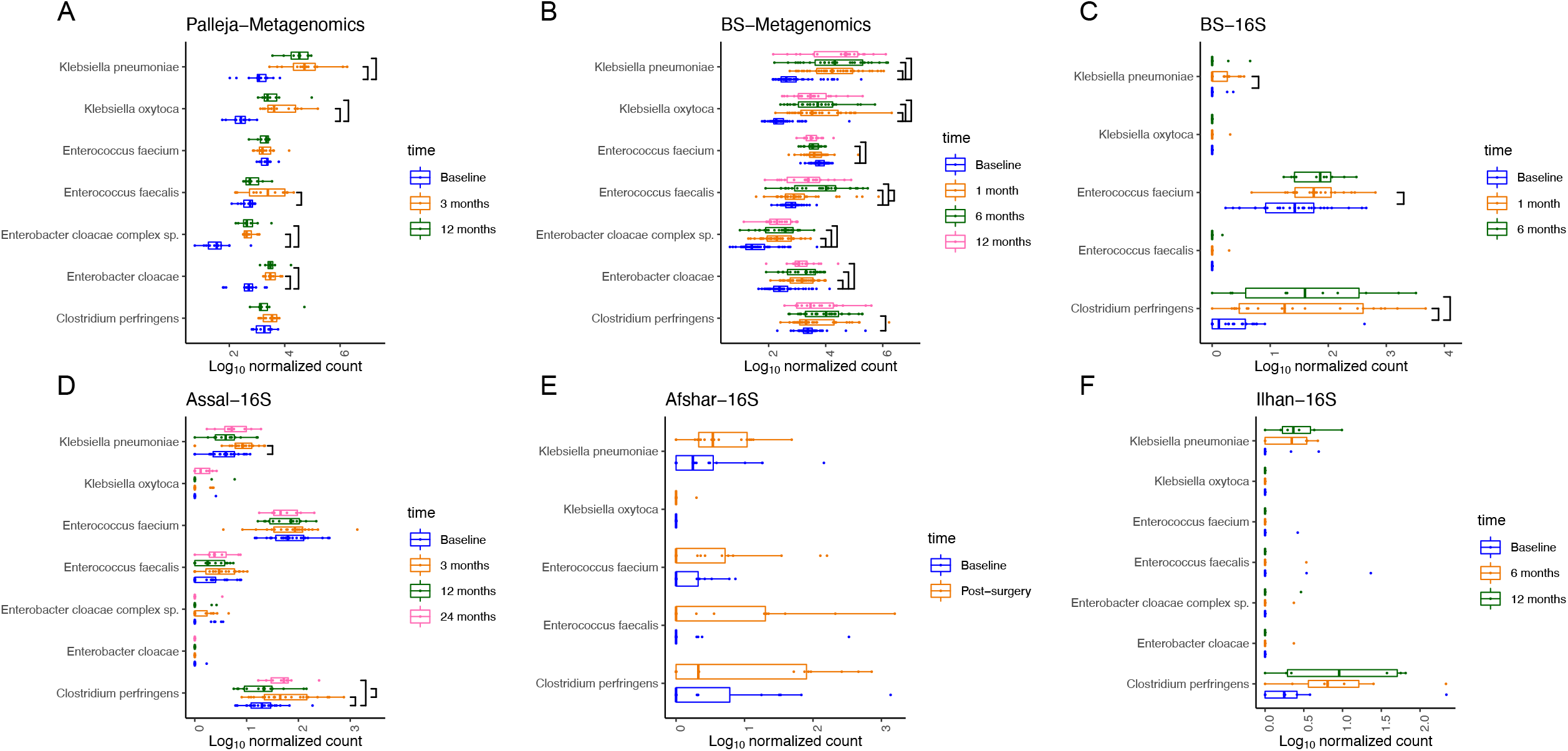
Roux-en-Y Gastric Bypass is Associated with Colonization of Opportunistic Pathogens in the Gut. Boxplot shows the log_10_ normalized count of opportunistic pathogens (*Klebsiella pneumoniae, Klebsiella* oxytoca, *Enterococcus faecalis, Enterobacter cloacae complex sp, Enterobacter cloacae, Enterococcus faecium*, and *Clostridium perfringens)* at pre- and post- surgery from the Kraken2- taxonomically classified metagenomic and 16S rRNA gene sequences. Mixed linear models were used to compare the log_10_ normalized count of opportunistic pathogens between different timepoints. Significant differences at FDR 5% are shown.

A similar trend was observed in the 16S rRNA gene datasets; however, only the increase in *Klebsiella pneumoniae* (BS study adjusted p = 0.007 at one month post-surgery; Assal study adjusted p = 0.04 at three months post-surgery) and *Clostridium perfringens* (BS study adjusted p < 0.001 at one and six months post-surgery and Assal study adjusted p = 0.002 at three months and adjusted p = 0.04 at two years after surgery) reached statistical significance (**Figure 5 C and D)**. This could be simply because Kraken2 may not have enough information to resolve many 16S rRNA gene sequences to this level of taxonomic assignment in many instances. Nevertheless, the increased trend in opportunistic pathogens post-surgery observed in both 16S rRNA gene and metagenomic datasets indicate that RYGB is associated with colonization of some of the opportunistic pathogens in the gut, which may remain abundant months after surgery.

## Discussion

Previous studies have shown that bariatric surgery induces significant shifts in the gut microbiota [2]. A recent systematic review of longitudinal bariatric surgery studies reported that similar patterns in microbial profiles were observed after RYGB and SG across studies [14]. For example, at the phylum level, *Proteobacteria* were increased after RYGB in numerous studies [3, 4, 10, 34]. In addition, at the genus level, *Prevotella [8, 35]* and *Viellonella [3, 34, 36]* were increased following RYGB in multiple studies. Despite these similarities, substantial inconsistencies in microbial profiles following surgery have also been observed across different cohorts. For example, there are conflicting results regarding changes in the relative abundance of the phylum *Bacteroidetes* as well as *Faecalibacterium* and *Bifidobacterium* species following bariatric surgery [3, 6-13]. In addition, some studies have reported an increase in microbial diversity post-surgery [5, 11, 12], while Paganelli et al. 2019 observed a short-term decrease in microbial diversity [37]. These conflicting results can be attributed to large variations in study designs, data collection methods, and data analyses across studies [14]. Factors such as diet, medications, physical activity, and disease states are not always well controlled across clinical studies and cannot necessarily be corrected in a post-hoc analysis. However, if sequence data for these studies are publicly available, it is possible to eliminate analytical differences between studies. The results we report here remove variations in data analysis between studies by applying identical bioinformatic tools and statistical models.

Our results revealed a significant and surprisingly robust microbial signature in the weeks and months following RYGB. By integrating multiple studies, our results confirm and expand on some of the original observations made in previous publications. For example, when we compared changes in taxa across two of 16S rRNA gene datasets (Assal and BS datasets), we found over 20 taxa that were significantly changed in both datasets with an un-adjusted p <0.05 and a remarkable degree of similarity in the magnitude of the changes across the two cohorts (Fig. 1C). By contrast, in the original paper, Assal et. al reported only ten taxa by Mann– Whitney paired test that were significantly changed. Therefore, our integrated analysis allowed us to achieve a better resolution of post-surgical changes in the microbial profile in a way that an individual study with a small sample size would be underpowered to detect.

In our analysis, we observed an increase in *Veillonella* and *Streptococcus* and a decrease in the *Blautia* (all from the Firmicutes phylum) across the four studies. These changes could have important clinical implications post-surgery. For example, *Veillonella* and *Streptococcus* metabolize lactate [38, 39], which consequently impacts butyrate metabolism [40, 41] and the integrity of the epithelial barrier [42]. Enhanced integrity of intestinal epithelium could decrease low-grade systemic inflammation and improve metabolic disorders [43]. Our study also revealed an in increase in *Akkermensia* within the *Verrucomicrobia* phylum. *Akkermensia* contains mucin degrading microbes and has been shown to increase after bariatric surgery in several studies [3, 12, 36, 44]. Previous animal studies have shown that *Akkermensia muciniphila* protects against obesity and diabetes through enhancing the intestinal epithelium barrier and potentially decreasing endotoxemia and low-grade inflammation [45, 46]. *Akkermensia muciphilia* was also associated with improvements in insulin sensitivity markers in humans [47]. We also observed that *Escherichia* (found in the Proteobacteria phylum) was increased, which is in agreement with previous studies [8, 11] and a negative correlation between *E. coli* and serum leptin after RYGB was previously reported [8].

In addition to the four 16S rRNA gene datasets, we compared our novel metagenomic sequences with the previously published Palleja metagenomic study. This allowed us to examine the effect of RYGB on the function of the gut microbiome in addition to the taxonomic composition. Our results showed a more robust and replicable signature in metabolic pathways following surgery across the two cohorts compared to the taxonomic profile. Changes in carbohydrate, amino acid, and lipid metabolism by the gut microbiome post-surgery could be due to dietary changes, the adaptation of the gut microbiome to the new gastrointestinal environment, or the increased potential of the gut microbiome in energy harvest as a compensatory response to the reduced food intake after RYGB [12].

Moreover, our study revealed that some opportunistic pathogens associated with the hospital environment, such as *Klebsiella pneumonia* and *Clostridium perfringens* increased after RYGB. Changes in the gastrointestinal environment and routinely administered prophylactic antibiotics prior to surgery might be the reasons for the increase in opportunistic pathogens after surgery. Future metagenomic studies are warranted to confirm these results and to determine the extent to which these opportunistic pathogens persist long-term after surgery and whether they have any effect weight outcomes and metabolic response to bariatric surgery.

This study has some limitations that should be noted. First, due to lack of availability of weight and metabolic data in some studies, we were unable to perform any association studies between the microbial signature with weight and metabolic outcomes after RYGB. Therefore, future studies are needed to characterize the effects of the taxa that were found commonly increased or decreased across studies on the outcomes of surgery. Second, again due to lack of data availability, we were not able to control for BMI, age, medications, diet, metabolic disorders and other covariates that could potentially impact gut microbiota composition. However, this limitation is less likely to have impacted our main findings since our outcomes show consistency across multiple studies. Finally, the microbial signature observed in this study might only reflect the immediate impact of surgery-related procedures which could include liquid diet or exposure to antibiotics. Future research is needed to determine if this consistent microbial signature persists over a longer term.

## Conclusions

In summary, our study highlights a robust signature in both microbial composition and gene function following RYGB across different cohorts. On-going assessment of our cohort will allow us to determine in future studies if this microbial signature is predictive of weight outcomes after surgery. These studies may allow us to use the microbial community to guide decisions on which subset of patients will successfully respond to surgery. Our characterization of the robust effect of the microbial signature across cohorts is a necessary pre-requisite for development of novel microbiome-based interventions for personalized treatment of obesity improving outcomes following surgery.

## Data Availability

Sequences have been deposited at in the NCBI SRA (National Center for Biotechnology information Sequence Read Archive (https://www.ncbi.nlm.nih.gov/sra) under BiopProject PRJNA668357 and PRJNA668472. Sequences from other datasets that are included in this study can be found at SRA under BioProject PRJNA395660, PRJNA612042, and PRJEB14082, PRJEB12947. Scripts and materials to reproduce all figures and tables are available at https://github.com/FarnazFouladi/RYGB_IntegratedAnalysis2020.

## List of abbreviations

RYGB: Roux-en-Y Gastric bypass
SG: Sleeve Gastrectomy
ASVs: Amplicon Sequence Variants
FDR: False Discovery Rate
LASSO: Least Absolute Shrinkage and Selection Operator
AUROC: Area Under the Receiver Operating Characteristic Curve
LOSO: Leave One Study Out (LOSO)

## Ethics approval and consent to participate

This study was approved by the Institutional Review Boards (Cleveland Clinic: #16-1460, Sanford Health: #00001409 and North Dakota State University #PH17112) at both data collection sites and all patients provided written informed consent prior to enrollment. This study is registered at www.clinicaltrials.gov with trial ID NCT03065426.

## Authors’ contributions

FF analyzed the data and wrote the manuscript, IMC contributed to 16S rRNA gene and metagenomic sequencing and to the review and editing of the manuscript, TJS provided statistical input and contributed to the review and editing of the manuscript, EBS contributed to 16S rRNA gene and metagenomic sequencing, LH (co-Principal Investigator) designed and conducted the clinical study, obtained funding, and contributed to the review and editing of the manuscript, KS (co-Principal Investigator) designed and conducted the clinical study, obtained funding, and contributed to the review and editing of the manuscript, AAF supervised data analysis and contributed to the review and editing of the manuscript.

