## Supplementary figures and images for "A Microbial Signature Following Bariatric Surgery is Robustly Consistent Across Multiple Cohorts"

### Supplementary Figure 1

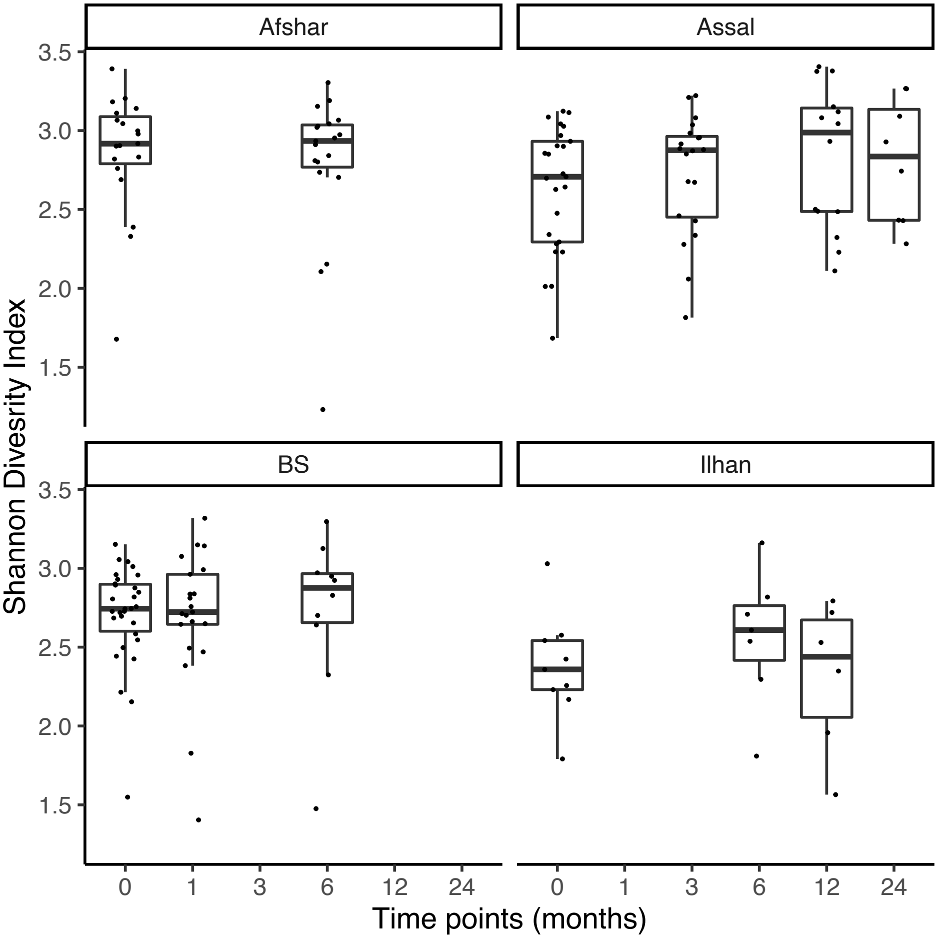

### Supplementary Figure 2

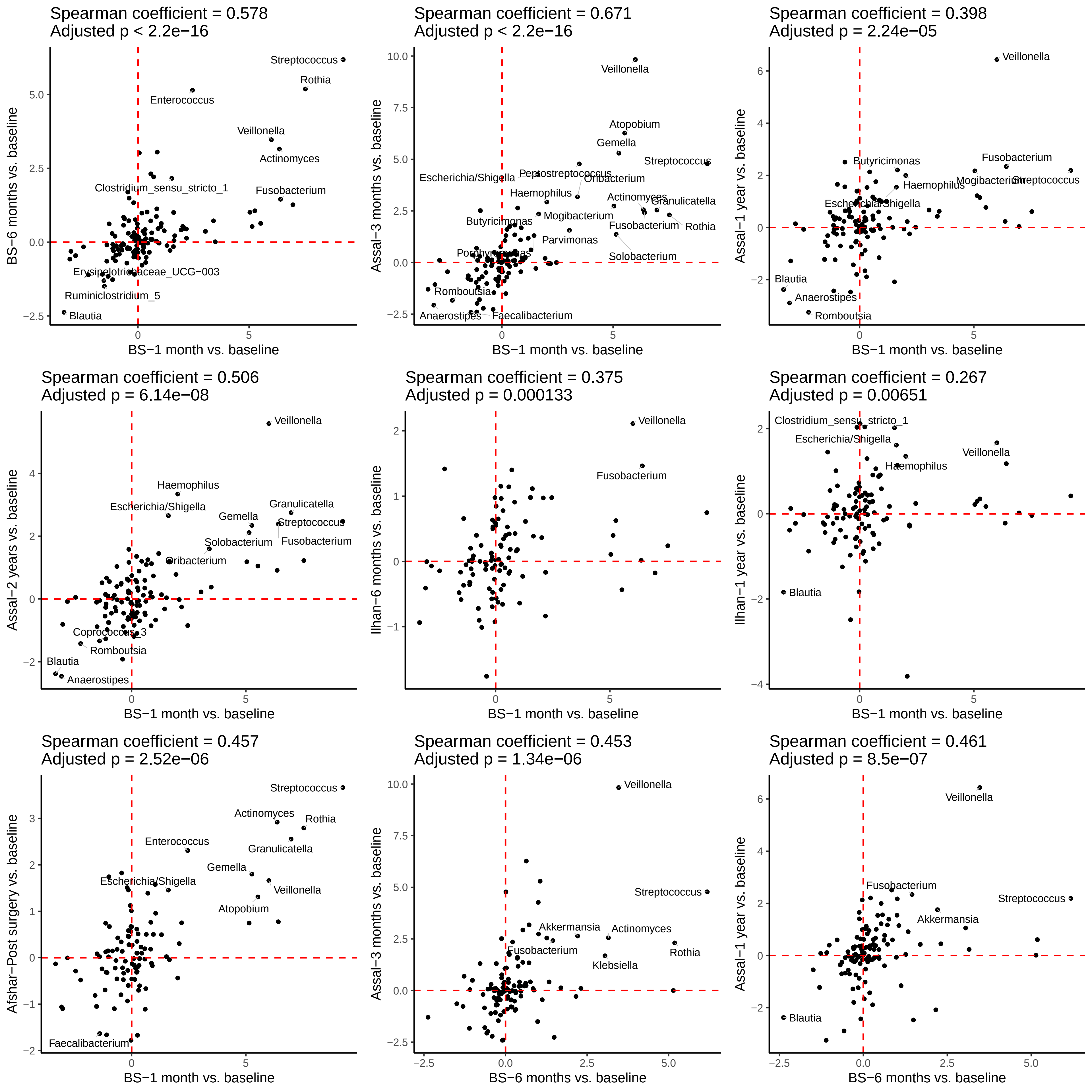

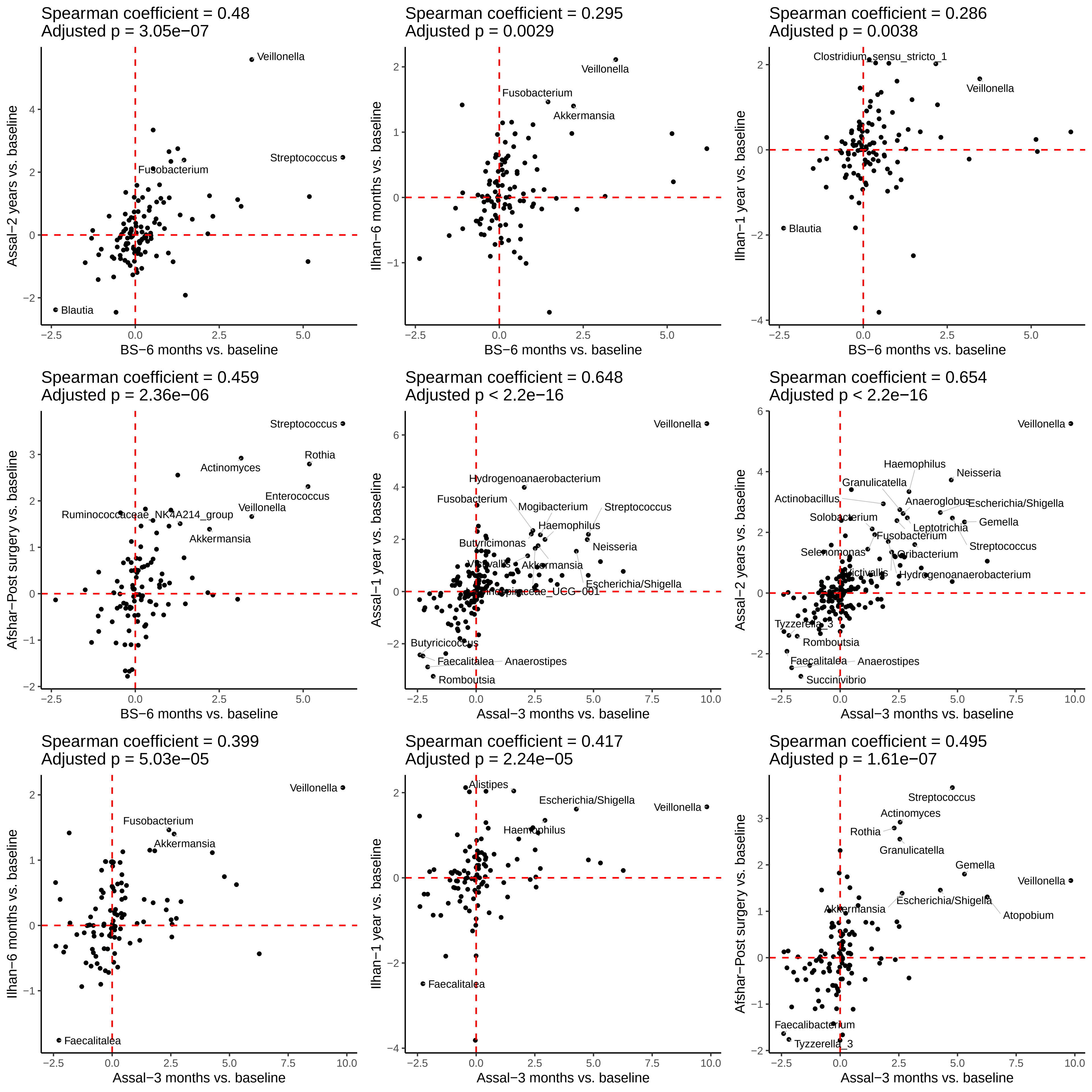

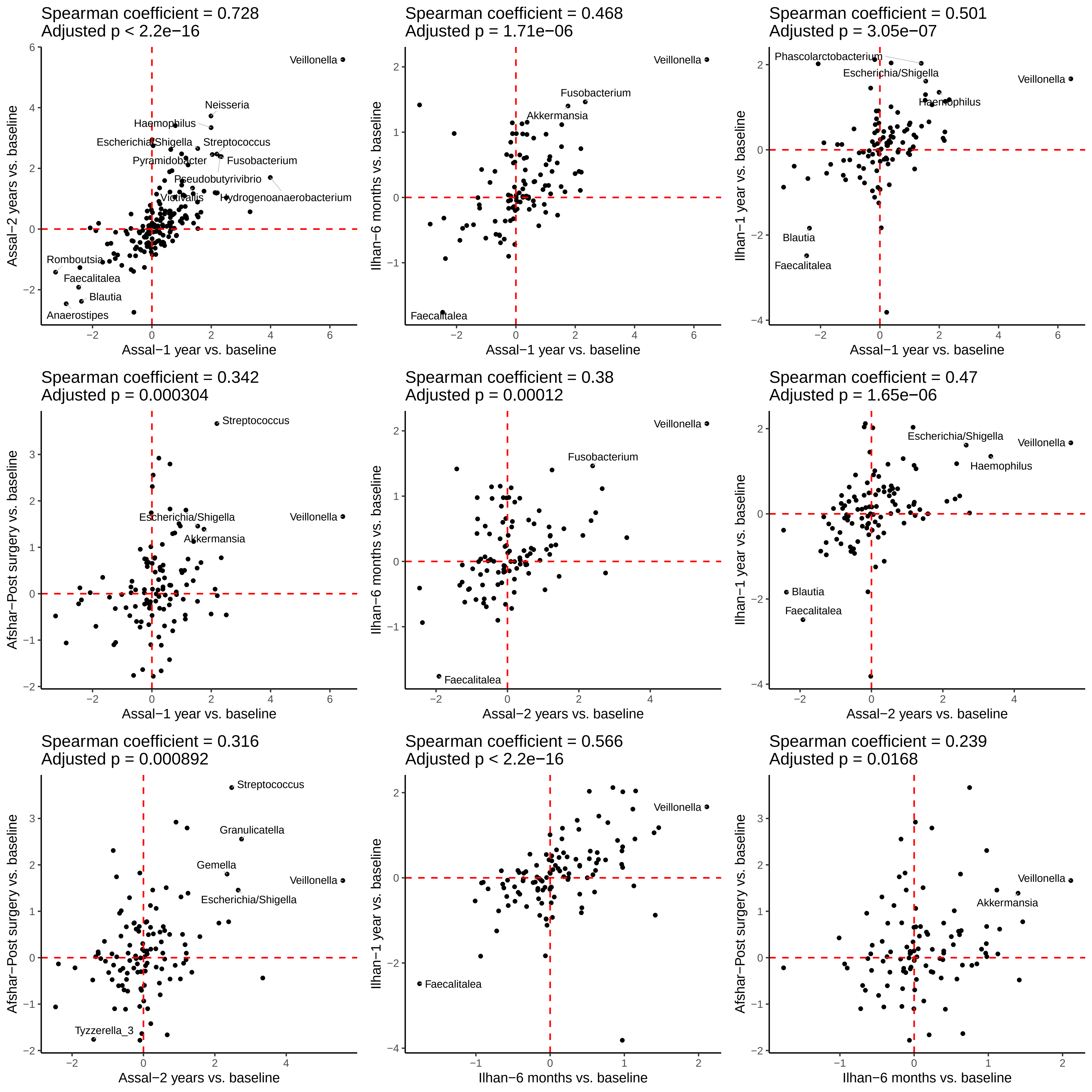

Spearman coefficient = 0.288
Adjusted p = 0.00404

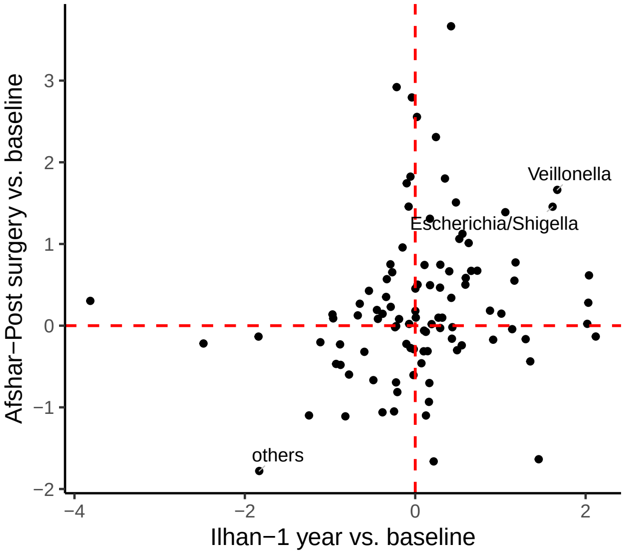

### Supplementary Figure 3

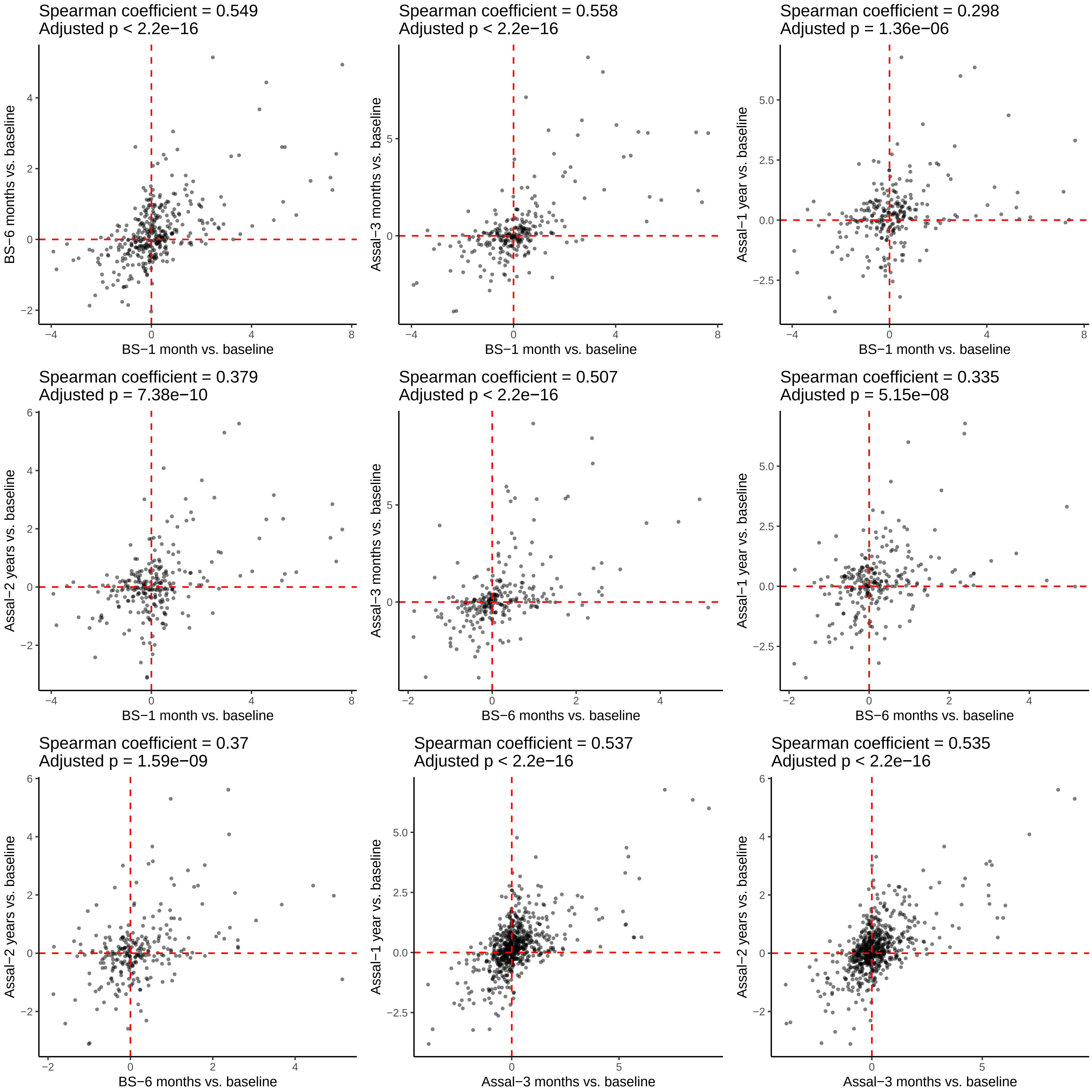

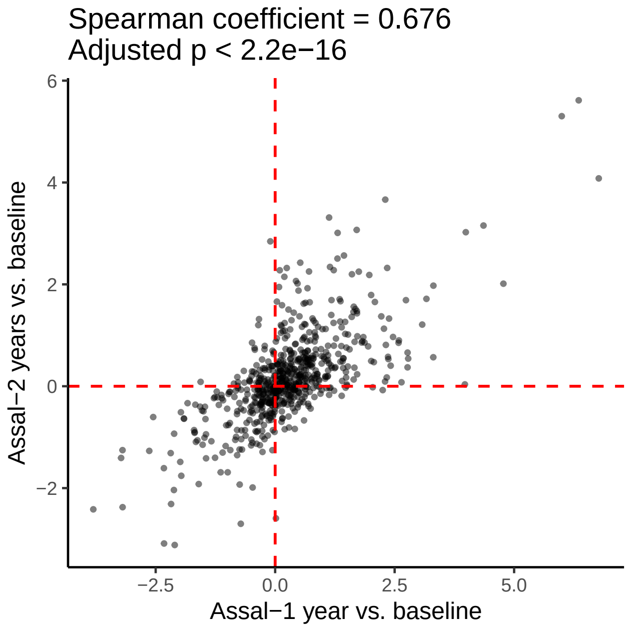

### Supplementary Figure 4

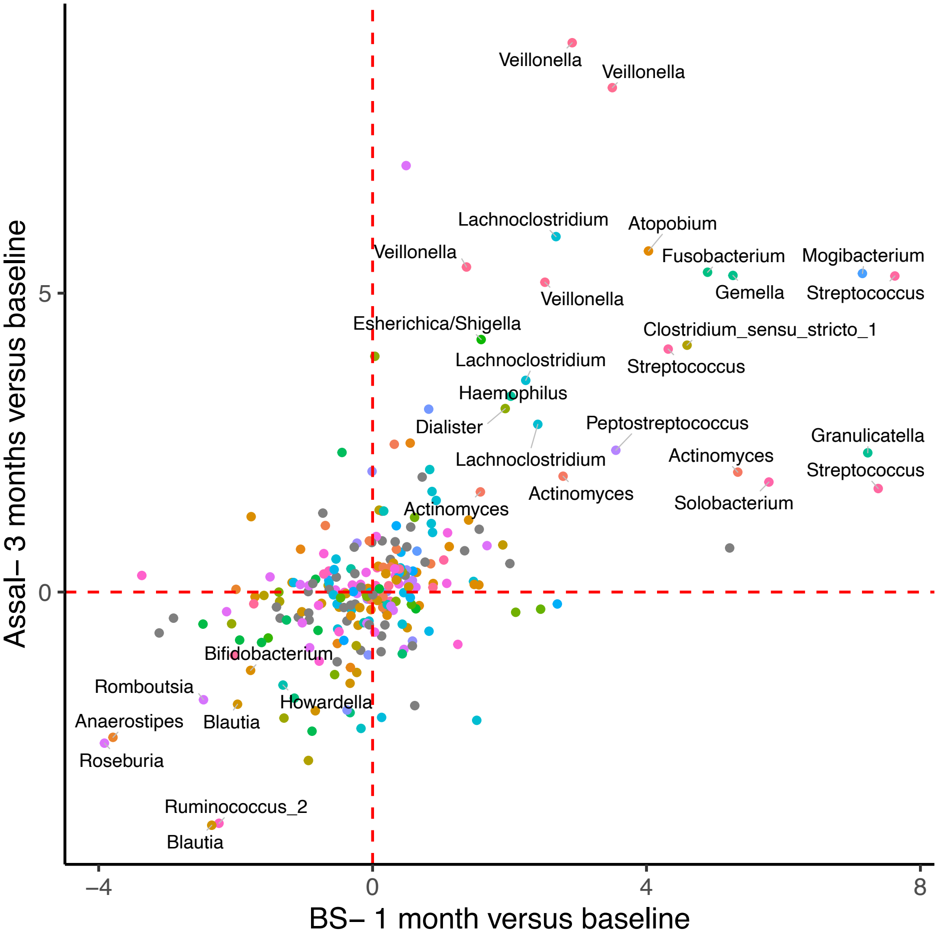

### Supplementary Figure 5

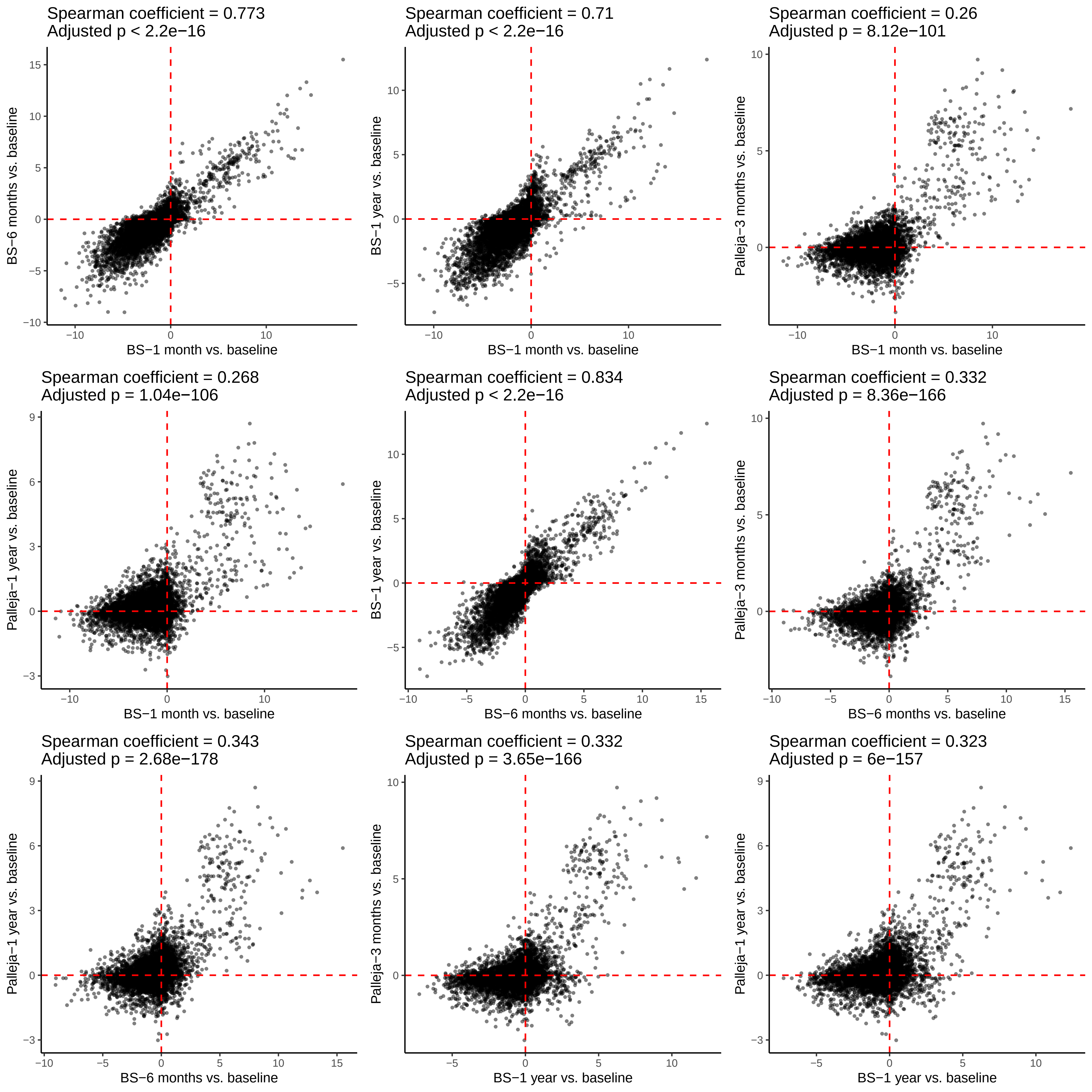

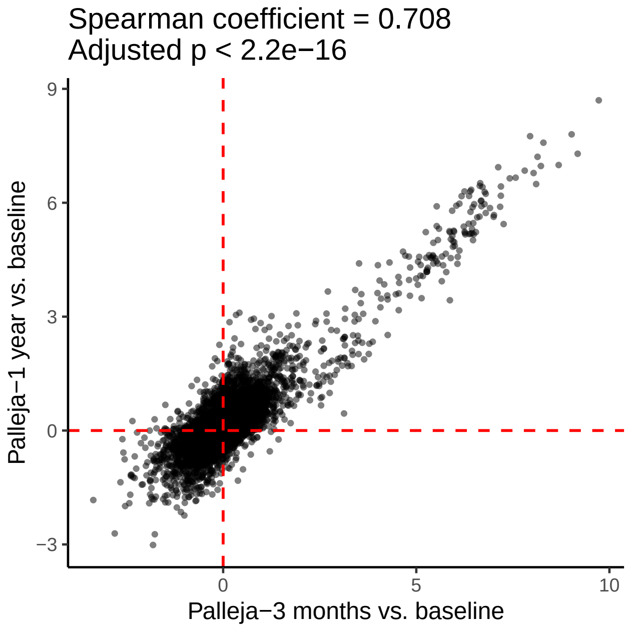

### Supplementary Figure 6

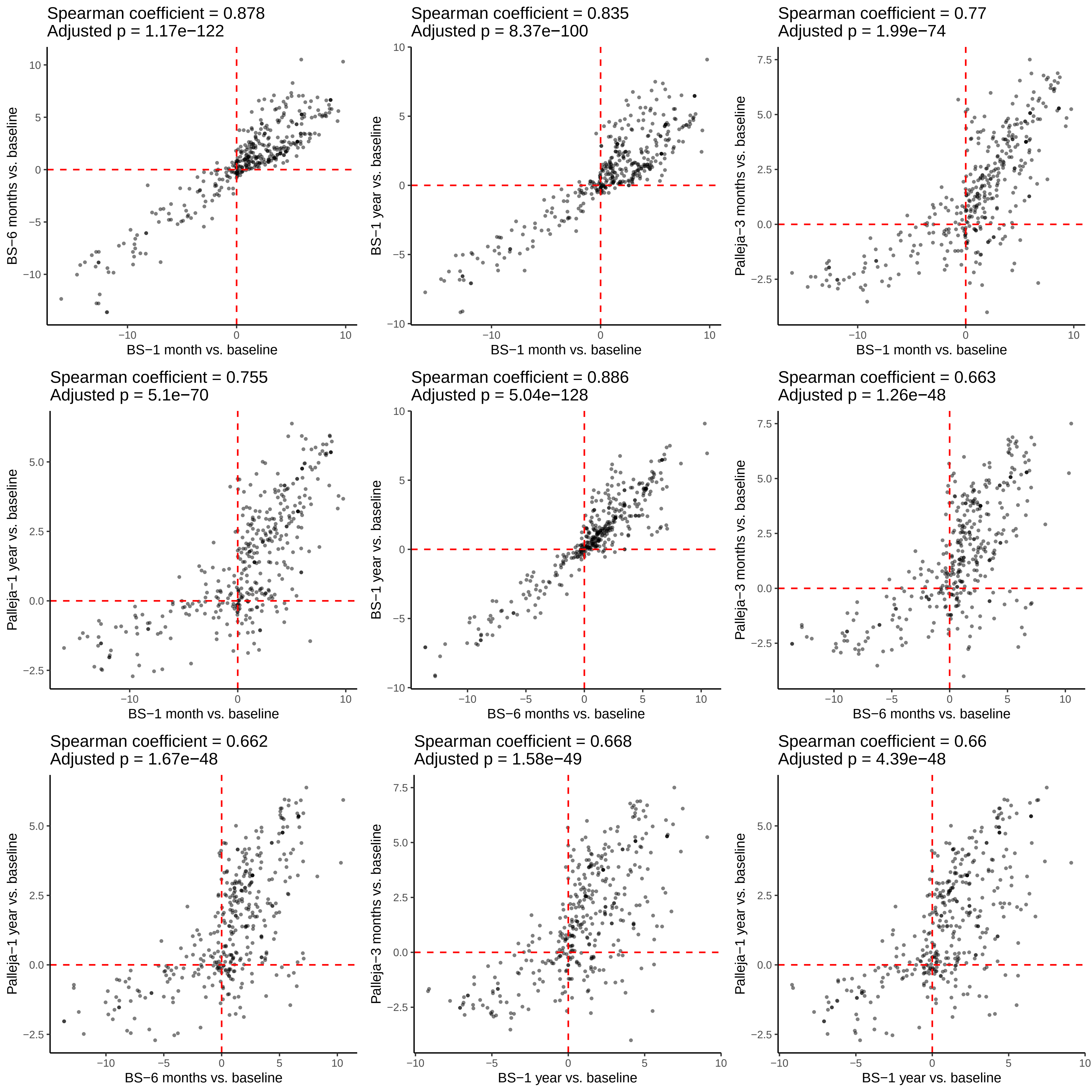

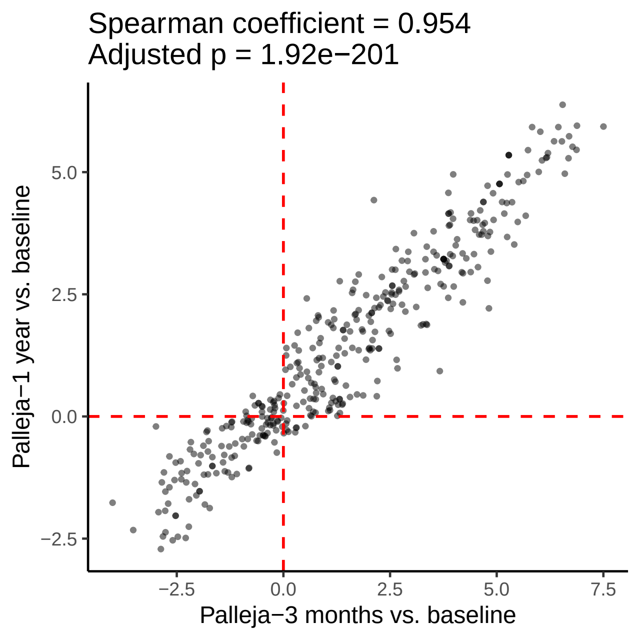

### Supplementary Figure 7

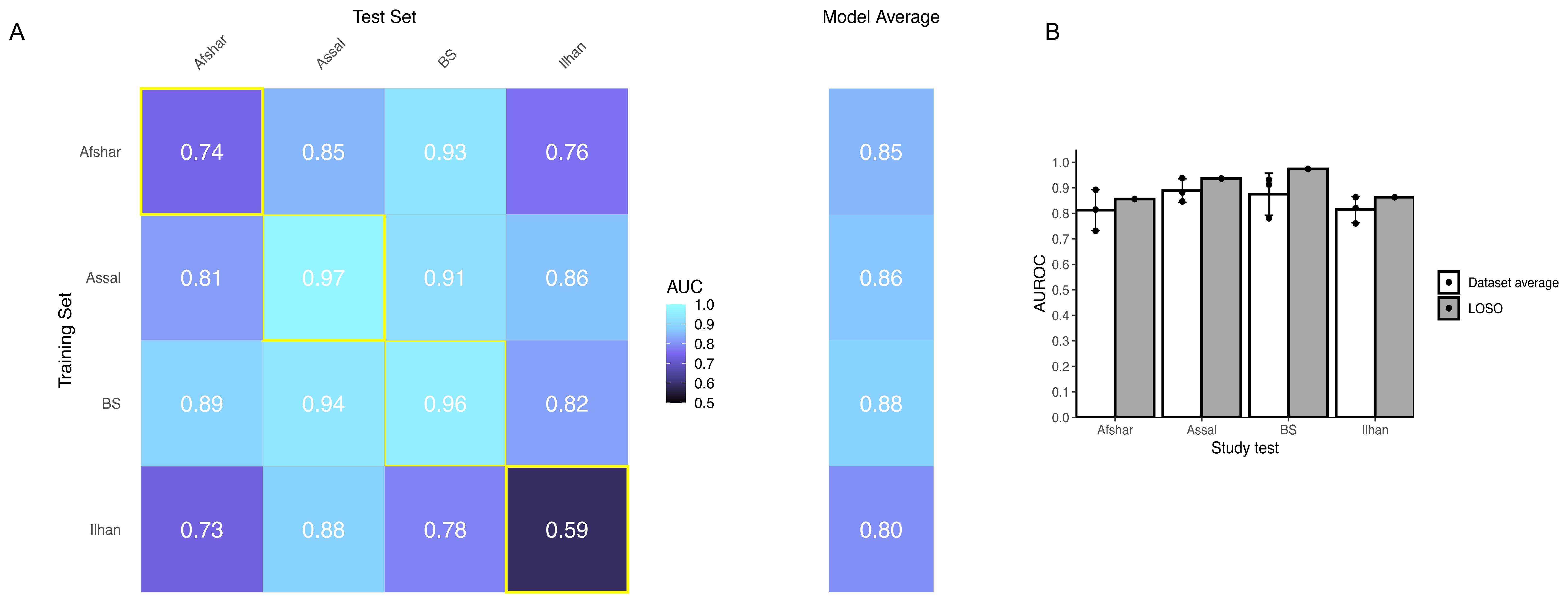
