## Supplementary Table and Figure Legends for "A Microbial Signature Following Bariatric Surgery is Robustly Consistent Across Multiple Cohorts"

**Legends for Supplementary Figures and Tables:**

**Supplementary Figure 1. Shannon Diversity Index Does Not Change Following Roux-en-Y Gastric Bypass Surgery in Four 16S rRNA Datasets.** Mixed linear models with timepoints as fixed effect and patient ID as random effects were used to compare Shannon Diversity Index at different timepoints.

**Supplementary Figure 2. A Consistent Signature of Fecal Microbiota at the Genus Level is Associated with Roux-en-Y Gastric Bypass Surgery Across 16S rRNA Studies.** Log_10_ p-value versus log_10_ p-value plots are generated using the p-values from mixed linear comparing log_10_ normalized count of taxa at each timepoint compared to baseline with patient ID as random effects. Upper right-quadrant and lower left-quadrant show taxa that were increased and decreased, respectively, in two different studies or at two different timepoints within a study. Spearman rank-order correlation was used to test the consistency of changes in taxa after RYGB between studies or within studies. Taxa with unadjusted p < 0.05 in two studies or at two timepoints within a study are annotated.

**Supplementary Figure 3. A Consistent Signature of Fecal Microbiota at the Sequence Variant Level is Associated with Roux-en-Y Gastric Bypass Surgery Across 16S rRNA Studies.** Log_10_ p-value versus log_10_ p-value plots are generated using the p-values from mixed linear models comparing log_10_ normalized count of sequence variants at each timepoint compared to baseline with patient ID as random effects. Upper right-quadrant and lower left-quadrant show sequence variants that were increased and decreased, respectively, in two different studies or at two different timepoints within a study. Spearman rank-order correlation was used to test the consistency of changes in sequence variants after RYGB between studies or within studies. Sequence variants with unadjusted p < 0.05 in two studies or at two timepoints within a study are annotated.

**Supplementary Figure 4. Sequence Variants that are Consistently Increased or Decreased in Both BS and Assal Datasets Belong to Different Genera.** Log_10_ p-value versus log_10_ p-value plot is generated using the unadjusted p-values from mixed linear models comparing the log_10_ normalized count of sequence variants at 1 month for the BS study and 3 months for the Assal study compared to baseline. Sequence variants that are increased or decreased in both studies are annotated.

**Supplementary Figure 5. A Consistent Signature of Fecal Microbiota at the Species Level is Associated with Roux-en-Y Gastric Bypass Surgery Across Metagenomic Studies.** log_10_ p-value versus log_10_ p-value plots are generated using the unadjusted p-values from mixed linear models comparing log_10_ normalized count of species at each timepoint post-surgery to baseline. The upper right-quadrant and lower left-quadrant show species that were increased and decreased, respectively, in two different studies or at two different timepoints within a study. Spearman rank-order correlation was used to test the consistency of changes in species after RYGB between studies or within studies.

**Supplementary Figure 6. A Consistent Signature of Metabolic Pathways is Associated with Roux-en-Y Gastric Bypass Surgery Across Metagenomic Studies.** log_10_ p-value versus log_10_ p-value plots are generated using the unadjusted p-values from mixed linear models comparing normalized count of metabolic pathways at each timepoint post-surgery to baseline. The upper right-quadrant and lower left-quadrant show metabolic pathways that were increased and decreased, respectively, in two different studies or at two different timepoints within a study. Spearman rank-order correlation was used to test the consistency of changes in species after RYGB between studies or within studies.

**Supplementary Figure 7. Supervised Classification Using the Random Forest Reveals that the Fecal Microbiota can be Used to Discriminate Pre- and Post-Samples Across 16S rRNA Studies.** (A) The heatmap shows the area under the receiver operating characteristic curve (AUROC) from cross validations within each study (yellow boxes along diagonal) and study-to-study model transfer (external-validations off-diagonal). The last column shows the average AUROC for study-to-study predictions. (B) White bar plots show the AURC for a model trained on data from a single study predicting the test study on the X-axis. The bar height represents the average AURC for the 3 classifiers and the error bar represents the standard deviation. The grey bar plots show the AURC for a model trained on all studies but one (LOSO validation).

**Supplementary Table 1.** All Bioprojects that include Roux-en-Y gastric bypass clinical studies are listed. Only three 16S rRNA datasets and one metagenomic dataset (highlighted) that their sequences and metadata are publicly available were included in our comparative analysis.

**Supplementary Table 2.** Mixed linear models were used to compare metabolic pathways at each timepoint compared to baseline in WGS datasets (A:BS, B:Palleja).
